# High-Field 7T MRI in a drug-resistant paediatric epilepsy cohort: image comparison and radiological outcomes

**DOI:** 10.1101/2024.08.19.24312117

**Authors:** Katy Vecchiato, Chiara Casella, Ayse Sila Dokumaci, Olivia Carney, Jon O. Cleary, Pierluigi Di Ciò, Michela Cleri, Kathleen Colford, Rory J. Piper, Tomoki Arichi, Michael Eyre, Fraser Aitken, Raphael Tomi-Tricot, Tom Wilkinson, Colm J McGinnity, Siti N Yaakub, Sharon L. Giles, Shaihan Malik, Alexander Hammers, Philippa Bridgen, David W. Carmichael, Jonathan O’Muircheartaigh

**Affiliations:** Department for Forensic and Neurodevelopmental Sciences, Institute of Psychiatry, Psychology and Neuroscience, King’s College London, London, UK; Research Department of Early Life Imaging, School of Biomedical Engineering and Imaging Sciences, King’s College London, London, UK; School of Biomedical Engineering and Imaging Sciences, King’s College, London, UK; London Collaborative Ultra high field System (LoCUS), London, St Thomas’ Hospital, UK; Department of Radiology, Guy’s and St. Thomas’ NHS Foundation Trust, London, UK; Guys and St Thomas’ NHS Foundation Trust, Kings College London, London, UK; Developmental Neurosciences Research and Teaching Department, UCL Great Ormond Street Institute of Child Health, London, UK; Department of Neurosurgery, Great Ormond Street Hospital, London, UK; MRC Centre for Neurodevelopmental Disorders, King’s College London, UK; Children’s Neurosciences, Evelina London Children’s Hospital at Guy’s and St Thomas’ NHS Foundation Trust, London, UK; MR Research Collaborations, Siemens Healthcare Limited, Camberley, UK; King’s College London & Guy’s and St Thomas’ PET Centre, School of Biomedical Engineering and Imaging Sciences, King’s College London, London, UK

## Abstract

**Background and Objectives:** Epileptogenic lesions in focal epilepsy can be subtle or undetected on conventional brain MRI. Ultra-high field (7T) MRI offers higher spatial resolution, contrast and signal-to-noise ratio compared to conventional imaging systems and has shown promise in the presurgical evaluation of adult focal epilepsy. However, the utility of ultra-high field MRI in paediatric focal epilepsy, where malformations of cortical development are more common, is unclear. This study compared 7T to conventional 3T MRI in children with epilepsy by comparing: (i) scan tolerability; (ii) radiological image quality; (iii) lesion yield.

**Materials and Methods:** Children with drug-resistant focal epilepsy and healthy controls were recruited prospectively and imaged at both 3T and 7T. Safety and tolerability during scanning was assessed via a questionnaire. Image quality was evaluated by an expert paediatric neuroradiologist and estimated quantitatively by comparing cortical thickness between field strengths. To assess lesion detection yield of 7T MRI, a multi-disciplinary team jointly reviewed patients’ images.

**Results:** 41 patients (8-17 years, mean=12.6 years, 22 male) and 22 healthy controls (8-17 years, mean=11.7 years, 15 male) were recruited. All children completed the scan, with no significant adverse events. Higher discomfort due to dizziness was reported at 7T (p=0.02), with side-effects more frequently noted in younger children (p=0.02). However, both field strengths were generally well-tolerated and side-effects were transient. 7T images had increased inhomogeneity and artefacts compared to those obtained at 3T. Cortical thickness measurements were significantly thinner at 7T (p<0.001). 8/26 (31%) patients had new lesions identified at 7T which were not identified at 3T, influencing the surgical management in 4/26 (15%).

**Discussion:** 7T MRI in children with epilepsy is feasible, well-tolerated and is associated with a 31% improvement in lesion detection rates.

## Introduction

Epilepsy is a chronic neurological condition that profoundly impacts health and quality of life (1). Approximately 40% of patients with focal epilepsy are drug-resistant (2). In approximately two-thirds of patients, resection of epileptogenic tissue can lead to seizure-freedom, or a marked (>90%) reduction in seizures (3,4), as well as potential improvements in cognitive and developmental outcomes (5).

While the identification of a lesion on imaging is the single biggest predictor of surgical outcome (6–8), this can be challenging in children. Conventional (1.5T or 3T) brain MRI can be used to identify structural epileptogenic abnormalities, however about 30% of all focal epilepsy patients (both adults and children) present with no findings on MRI (MRI negative, 5–8). This is especially true for focal cortical dysplasias (FCDs) and other malformations of cortical development, which can be small and visually subtle in radiological appearance (13), and are more common in children (14). Failure to detect a structural lesion on MRI adds complexity to neurosurgical planning, can lead to further invasive investigations, and is associated with a less favourable prognosis (15).

Positron emission tomography (PET) can help localize occult focal abnormalities by highlighting alterations in focal metabolism (16,17), and its diagnostic sensitivity is further improved after co-registration with MRI (18). However, the area of hypometabolism identified with PET scans often extends beyond the epileptogenic zone, thus this technique cannot precisely define the lesion surgical margins (19).

Ultra-high field (7T) MRI offers higher signal to noise ratio and contrast compared to conventional MRI, enabling higher spatial resolution. This translates into a clearer delineation of anatomical structures, potentially increasing radiological detection and improving diagnostic confidence (20). Previous studies have demonstrated that 7T MRI significantly improves the detection and clinical decision-making for epilepsy in adults, revealing additional lesions and enhancing diagnostic confidence, with a diagnostic gain over conventional MRI ranging between 22% and 43% (21–25). However, the clinical efficacy of 7T MRI for lesion detection in children is not known. Moreover, there is no direct comparison of the performance of 7T MRI with clinically available 3T MR imaging in this clinical population.

Importantly, a comprehensive understanding of the practical utility of 7T MRI in children hinges on recognizing its associated side effects and how it is perceived during examinations. While studies in adults have consistently demonstrated that discomfort is transient and 7T scans are generally well-tolerated (26,27), the experience may vary in children due to age-related factors and developmental stages.

In this work, we present findings from a large prospective 7T MRI paediatric case series with a paired 3T acquisition, to investigate the relative tolerability, image quality and clinical yield of 7T MRI in children with drug resistant focal epilepsy.

## Methods

### Participants

Patients were identified from epilepsy outpatient clinics at the Evelina London Children’s Hospital, King’s College Hospital and Great Ormond Street Hospital. Healthy controls were recruited from existing volunteer databases, mainstream schools, and from the King’s College London recruiting webpage. Inclusion and exclusion criteria are summarised in Figure 1. PET scans were available in 29/41 patients, which were co-registered with the MRI images as part of clinical reporting (28), however this was not an explicit inclusion criterion.

**Figure 1.**
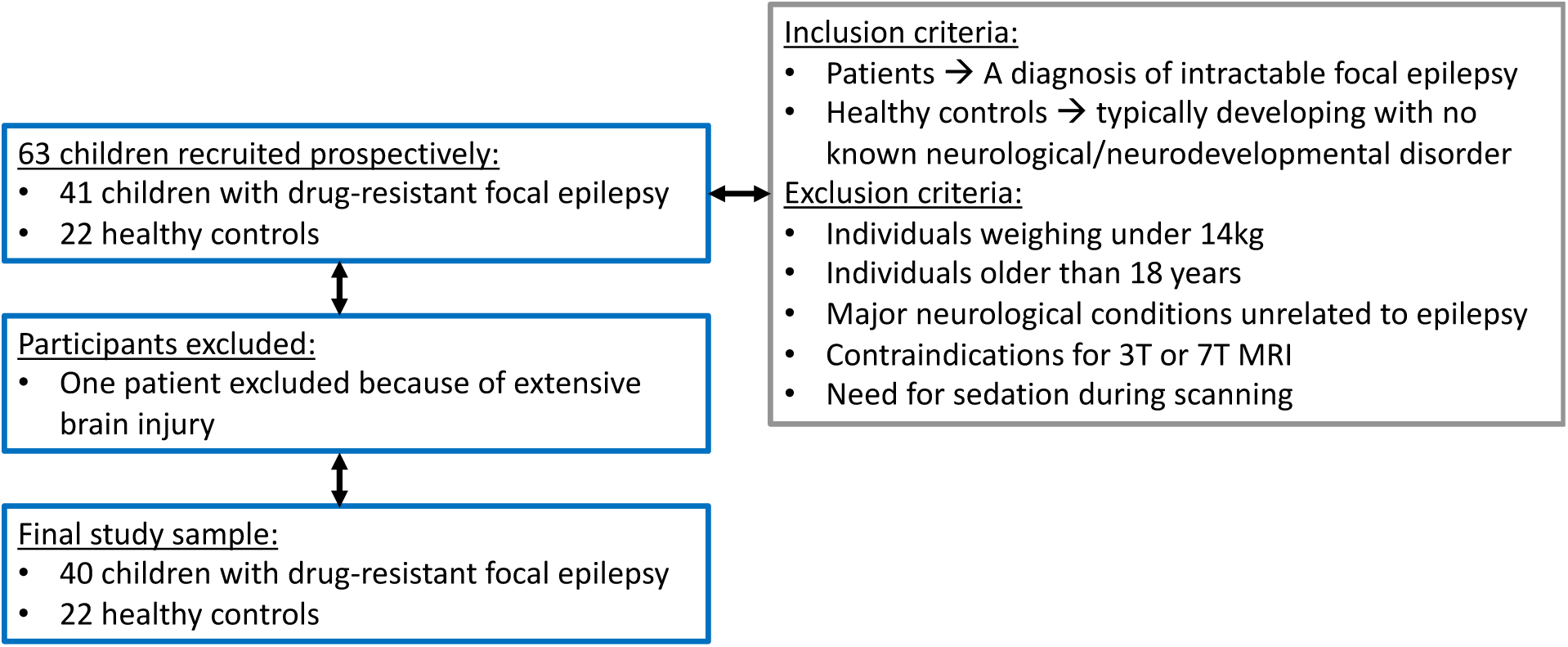
Flow diagram. Inclusion/exclusion criteria, initial number of participants and those excluded from the study are shown. Intractable = failure of adequate trials of two tolerated and appropriately chosen antiepileptic drugs.

### Clinical Background

The clinical details of the patients recruited are summarized in Supplementary Table 1. In addition to previous MRI data, patients’ clinical information and previous investigations were accessible.

### Data Acquisition

All participants undertook a 3T MRI including MPRAGE, FLAIR and T2-weighted sequences. Scans were acquired using the DISORDER (Distributed and incoherent sample orders for reconstruction deblurring using encoding redundancy) scheme (29), which has demonstrated improved tolerance against motion (30). In-depth detail of the reconstruction algorithm has been described previously (29,30). Participants also undertook a 7T MRI including 3D MP2RAGE (31), 3D FLAIR and 2D turbo spin-echo (TSE) T2-weighted sequences (32). Acquisition parameters are shown in Table 1.

**Table 1.** Acquisition details.

| Scanner | Sequence | TR/TE | Resolution | Acquisition Time |
| --- | --- | --- | --- | --- |
| **3T** | MPRAGE | 7.7/3.6ms | 1mm isotropic | 4min 46s |
|  | FLAIR | 5000/422ms | 1mm isotropic | 8 min 30s |
|  | T2-weighted | 2500/344ms | 1mm isotropic | 5 min 42s |
| **7T** | MP2RAGE (31) | 4000/3.15ms | 0.65mm isotropic | 7 min 18s |
|  | 1Tx FLAIR | 9000/240ms | 0.8mm isotropic | 7 min 5s |
|  | PTx FLAIR | 9000/325ms | 0.8mm isotropic | 6 min 54s |
|  | 2D TSE T2-weighted<br>(axial, coronal oblique,<br>sagittal)(32) | 8100/82ms | 0.4 x 0.4 x 2.0mm | 4 min 5s each |

Abbreviations: MPRAGE: Magnetization Prepared Rapid Gradient Echo Imaging; FLAIR: Fluid attenuated inversion recovery; MP2RAGE: Magnetization Prepared 2 Rapid Acquisition Gradient Echoes; 1Tx: single transmit; PTx: parallel transmit.

The 3T scanning session was carried out at the Evelina Neuroimaging Centre on an Achieva 3.0T system (Philips Healthcare, Best, The Netherlands), while the 7T scan was performed in the London Collaborative Ultra-High-Field System (LoCUS) facility on a MAGNETOM Terra system (Siemens Healthineers, Erlangen, Germany). Both facilities are located at St Thomas’ Hospital. The two scanning sessions were performed with a maximum interscan interval of 4 months. All participants were unsedated, wore earplugs for hearing protection, and watched a movie during scanning. Acquisition details are described in Table 3.

After the 21^st^ participant, the 7T MRI protocol was adapted so that the FLAIR sequence was acquired with a parallel radiofrequency (RF) transmission (PTx) coil with the system switched to prototype research configuration, to correct for B_1_ field inhomogeneities affecting the right temporal lobe (Figure 2A). When scanning in PTx mode, the FLAIR sequence from the PASTeUR package (33) was used in 3D, as well as the DiSCoVER method (32) for 2D T2-weighted imaging.

**Figure 2.**
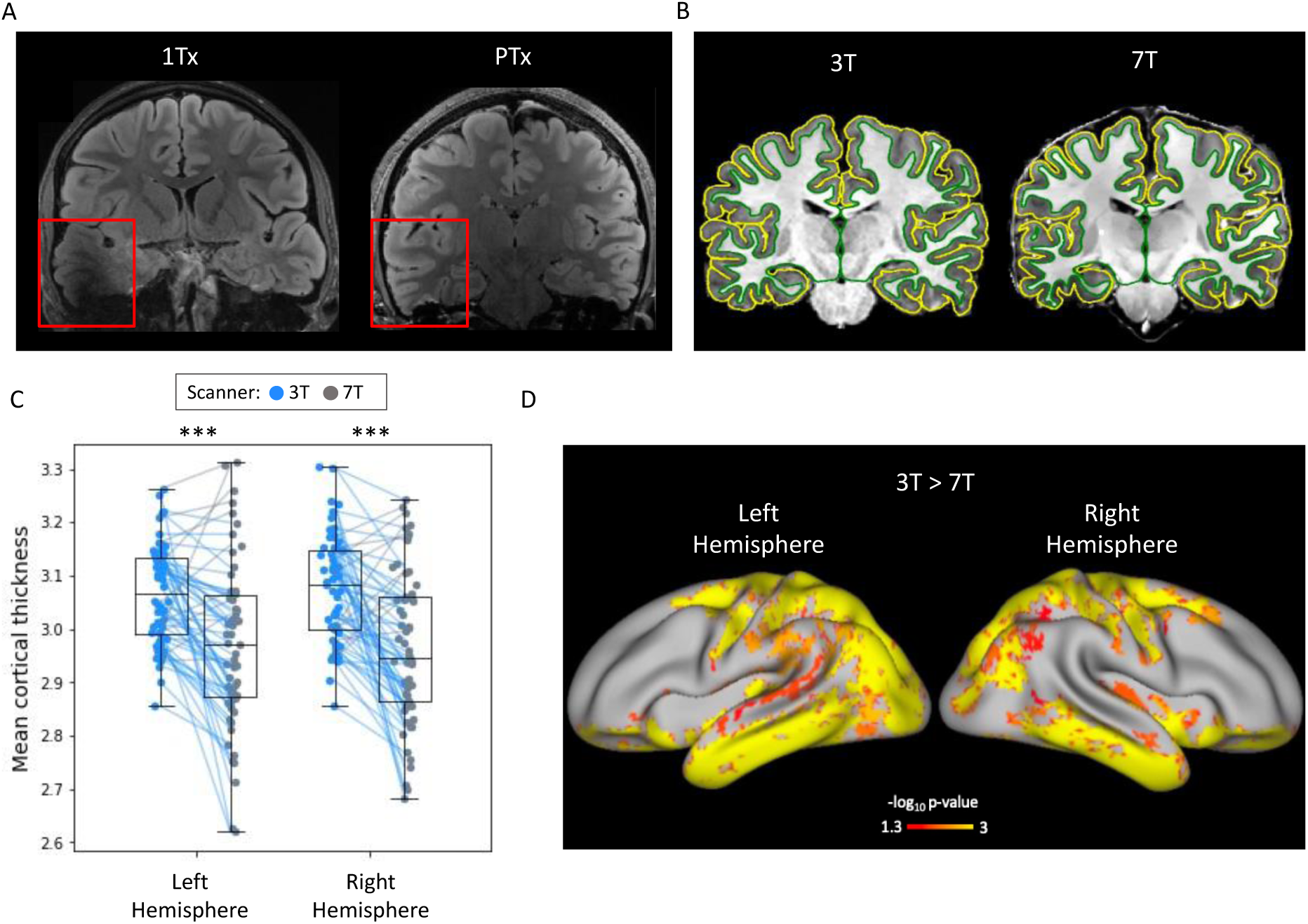
Imaging characteristics of 3T and 7T MRI. A) Impact of parallel RF transmission on FLAIR image of a healthy control. In A, acquired with single RF transmission (1Tx), there is a visible signal drop in the right temporal lobe. No signal drop is present in B, acquired with parallel transmit (PTx). B) Example of a single subject cortical delineation at 3T and 7T. C, D) Cortical Thickness Evaluation. Overall, the detected cortical thickness was thinner at 7T (C), with thinner boundaries in temporo-parietal/sensory regions (D).

### Analysis

#### Safety and tolerability

A survey adapted from that of Chou et al. (34) was employed to investigate general comfort and the presence of temporary collateral effects during 7T scanning in a subgroup of 45 participants (patients and controls) (see Supplementary Figure 1). In particular, the survey focused on feelings of discomfort, dizziness, noise, metallic taste in the mouth and feelings of parts of the body twitching.

Patients aged 8–11 and 12–17 years were given age-appropriate questionnaires and asked to rate the answers according to a five-point Likert scale, while for those aged 5-7 years pictograms were used to explain those feelings and asked for ‘yes’ or ‘no’ answers. Overall, a total of 21 surveys were collected for the group aged 8-11 years, 20 for the group aged 12-17 years and 4 for the group aged 5-7 years.

A Mann-Whitney test was conducted to analyse differences in tolerability and collateral effects at 7T between the groups aged 8-11 and 12-17. The youngest cohort sample size was too small for formal statistical testing and was excluded from the analysis.

Additionally, to test differences in tolerability between 3T and 7T, matched questionnaires were collected in a subgroup of eleven 8-to 16-year-old participants (mean age=13). The answers were dichotomised into ‘yes’ (score 1-3) and ‘no’ (score 4,5). Differences were tested using McNemar’s χ2 test for dichotomised data.

#### Quantitative assessment of cortical thickness

Pre-processing steps for 3T images included motion-correction with DISORDER (29) and Gibbs-ringing-correction (35). 7T MR images were denoised (36) and bias-field-corrected (37). Both 3T and 7T images were then processed with the Human Connectome Project (HCP) structural pipeline to perform cortical surface reconstruction and cortical thickness computation (38) (Figure 2B). Both T1-weighted (3T: MPRAGE, 7T: denoised combined ‘UNI’ image from the MP2RAGE acquisition) and T2-weighted scans (3T, 7T: FLAIR) were used for segmentation, the latter specifically to improve pial surface reconstruction.

Mean cortical thickness was extracted for both hemispheres, and a within-subjects comparison was carried out between 3T and 7T, investigating regional brain areas in cortical thickness as a surrogate for improved delineation of the grey-white matter boundary.

Statistical testing was performed on the cortical surface-based measurements using paired t-tests, tested through permutation with the PALM (Permutation Analysis of Linear Models) analysis package (39). Threshold-free cluster enhancement (TFCE) was employed as a test statistic. Age and sex were included as covariates. Correction for family-wise error rate was carried out across contrasts (contrast 1: 7T>3T; contrast 2: 3T>7T).

#### Radiological image quality assessment

To assess image visual quality, an expert paediatric neuroradiologist not involved in the clinical assessment reviewed the first 20 participants who underwent the 7T MRI, using a rating scale from Wang et al. (40). This expert was blinded to any demographic data, group identity, electrophysiology data, and PET information. They evaluated both 3T and 7T MRI images -comparing 3T MPRAGE with 7T MP2RAGE for T1-weighted contrast and 3T FLAIR with 7T FLAIR for T2-weighted contrast. The assessment included a subgroup of 10 patients (a mix of lesion-positive and lesion-negative) and 10 healthy controls. For 3T images, only motion-corrected versions were shown. The scoring was based on the presence of artifacts that could affect image quality and readability, such as motion, pulsation, and inhomogeneity.

#### Clinical value of 7T MRI

3T and the 7T images were reviewed by three expert neuroradiologists with longstanding experience in paediatric epilepsy imaging, the neurology, neurosurgery and neurophysiology team and, where relevant, a PET expert. Consensus in imaging interpretation was based on the identification of a structural abnormality that was concordant in terms of focus location with electroclinical and, when available, PET data.

### Standard Protocol Approvals, Registrations, and Patient Consents

Ethical approval was granted by the UK Health Research Authority and Health and Care Research Wales (ethics ref. 18/LO/1766). Written signed consent was obtained from the parents (or the person with legal parental responsibility) prior to data collection.

### Data Availability

The clinical and neuroimaging data used in the current work are available from the senior author (J.O.M.) on formal request indicating name and affiliation of the researcher as well as a brief description of the intended use for the data. All requests will undergo King’s College London-regulated procedure, thus requiring submission of a Material Transfer Agreement. Full preprocessing steps and the code to run the HCP preprocessing pipeline can be found at https://github.com/Washington-University/HCPpipelines. Please also see https://github.com/Washington-University/workbench for the source code for Connectome Workbench. Other code excerpts, information regarding the analysis, or intermediary results can be made available upon request to.

## Results

### Participants

63 children were recruited: 41 with drug-resistant focal epilepsy [8-17 years, mean age=12.6 ± 2.4 years, 22 male] and 22 healthy controls [8-17 years, mean age=11.7 ± 2.7 years, 15 male]. 1 patient was excluded from the analysis because of widespread abnormalities due to extensive brain injury.

### Safety and tolerability

No adverse events occurred during the 3T or 7T MRI scans. The 7T scan was generally well-tolerated, with only mild and transient discomfort (related to neck pain, or ear pain due to headphones position) reported in 38% of participants. More uncommon were dizziness, sensation of a metallic taste in the mouth and body twitching, reported in 25% of 7T scans.

The McNemar’s χ² test for dichotomized data indicated a statistically significant increase in reports of dizziness at 7T, observed in 55% of participants, compared to 3T, where it was reported in only 9% of participants (p = 0.02). However, this effect was reported as temporary, and only occurred whilst the patient was moving in and out of the scanner bore on the bed.

The most common reported discomfort associated with the scans was noise, which was defined as loud by about 67% of the population. There was no difference in terms of noisiness, discomfort, metallic taste in the mouth and body twitches between field strengths.

The Mann-Whitney test conducted to analyse differences between the groups aged 8-11 and 12-17 in terms of 7T-associated effects, showed that noise and sensation of metallic taste in the mouth were both more common in the younger group (p=0.02).

### Quantitative assessment of cortical thickness

Mean cortical thickness was different between 3T and 7T for both right [3T: Mean=3.06mm, 95% CI=(3.05, 3.10); 7T: Mean=2.98mm, 95% CI=(2.93, 3.00)] and left [3T: Mean=3.06mm, 95% CI=(3.04, 3.09); 7T: Mean=2.96mm, 95% CI=(2.93, 3.01)] hemispheres, with lower mean values detected at 7T (p<0.001) (Figure 2C). The whole brain vertex-wise comparison revealed that cortical thickness was reduced at 7T compared to 3T in temporo-parietal/sensory regions (Figure 2D).

### Radiological image quality assessment

Image quality was rated as very good for 3T images with little or no artifacts reported in 90% of cases. At 7T, the MP2RAGE had little or no artifacts in 85% of cases, with moderate effects of motion in 15%. 7T FLAIR images in the scans evaluated for image quality were all acquired using single RF transmission. These images were all affected by a consistent signal loss in the right temporal lobe area due to the presence of B_1_ field inhomogeneities (Figure 2A): for this reason, 90% of images were scored as moderately artifacted, even if the majority of cortex was of high-quality. As shown in Figure 2A and 2B, parallel RF transmission significantly improves the signal loss in the right temporal lobe when single RF transmission is used. 10% of FLAIR images were also heavily affected by motion.

### Clinical value of 7T MRI

The review of the images of the 40 patients with epilepsy showed that lesion detection yield at 3T was 35% (14/40), which increased to 55% (22/40) at 7T. Therefore 8/26 3T lesion-negative patients had new findings when scanned at 7T. In 2 patients, these findings were non-specific (patient 2: ventricular asymmetry, left occipital horn more prominent than right; patient 24: asymmetrically deep sulcus in the left superior frontal gyrus). In the remaining 6 patients with new findings at 7T, 6 (100%) had PET findings (for 4/6 PET findings showed concordant localising features, while for 2/6 findings were not localising). Reviewing the first 5 cases that were 3T lesion-negative but lesion-positive at 7T, radiologists later confirmed that the lesion was retrospectively visible at 3T but was either ambiguous or was less conspicuous compared to 7T. Figure 3 summarizes results for patients overall (A) and for patients that underwent all three (3T MRI, 7T MRI, PET) imaging modalities (B).

**Figure 3.**
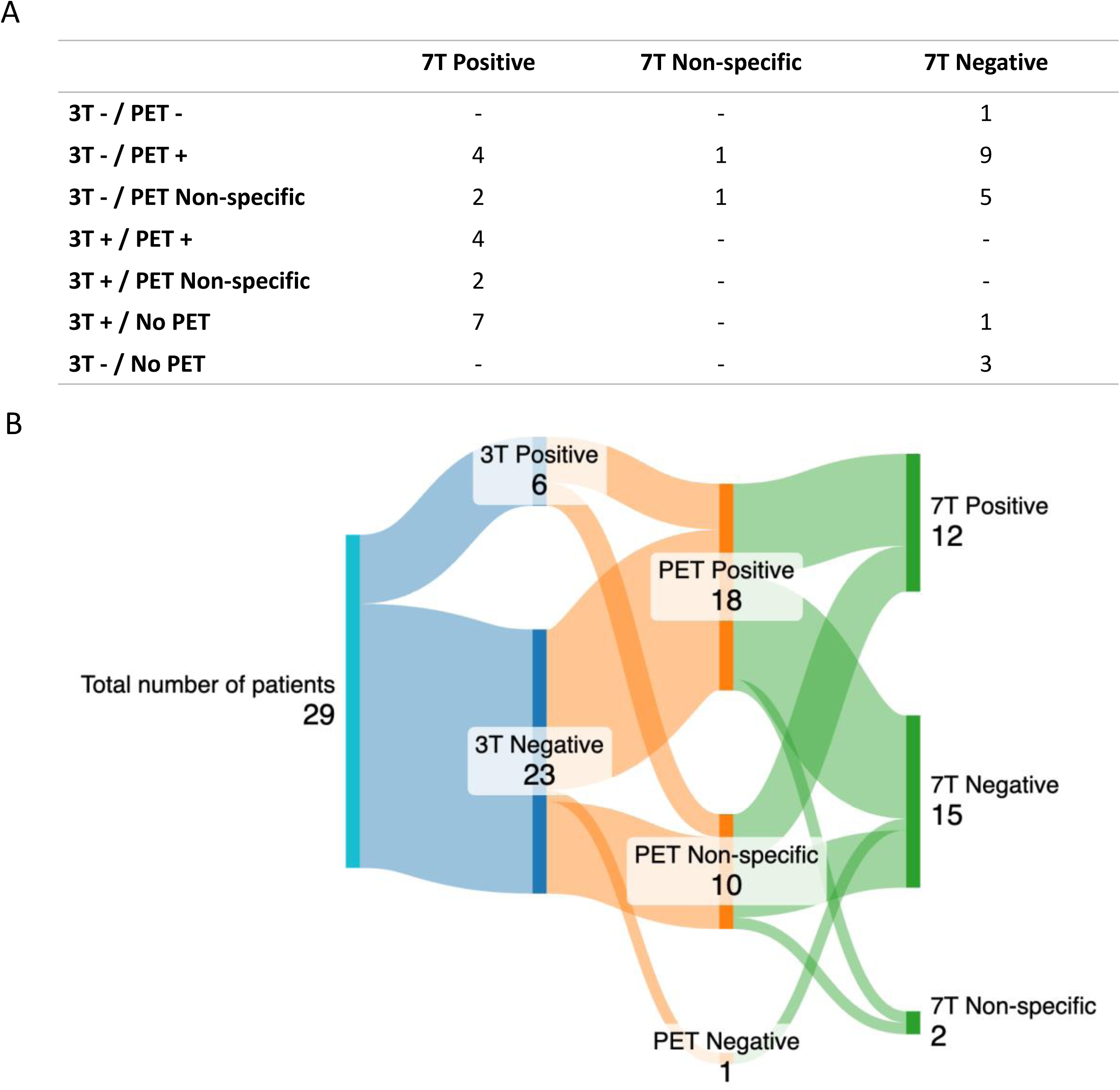
Lesion detection yield. A) Summary of findings across all patients. B) Sankey diagram of clinical outcomes in patients that underwent PET, 3T and 7T imaging.

Of the 8 patients with *new* findings at 7T, 2 (patient 1 and patient 7) underwent surgical resection and both were seizure-free at 3 years follow-up. As a result of the 7T findings, stereo-EEG was re-planned with a reduced number of electrodes in one patient and omitted in the other. In a further patient (patient 18), the SEEG demonstrated a focal onset of seizures in the putative lesion seen on the 7T MRI that was concordant with hypometabolism seen on the PET scan. Radiofrequency thermocoagulation was performed and the patient was seizure-free at 2 years follow-up. These three cases are described in detail in Figures 4, 5 and 6. For a fourth patient (patient 36), the 7T images helped with SEEG implantation and to better identify lesion margins. The lesion area detected with 7T MRI was consistent with the area detected with SEEG. For the remaining patients, the 7T scan findings did not impact clinical decision-making. Available clinical outcome details for all patients are reported in Supplementary Table 1. Patients 1, 5, 7, 18, 34 and 41 are those for which new specific findings were reported at 7T. As described above, for patients 2 and 24, new unspecific findings were reported.

**Figure 4:**
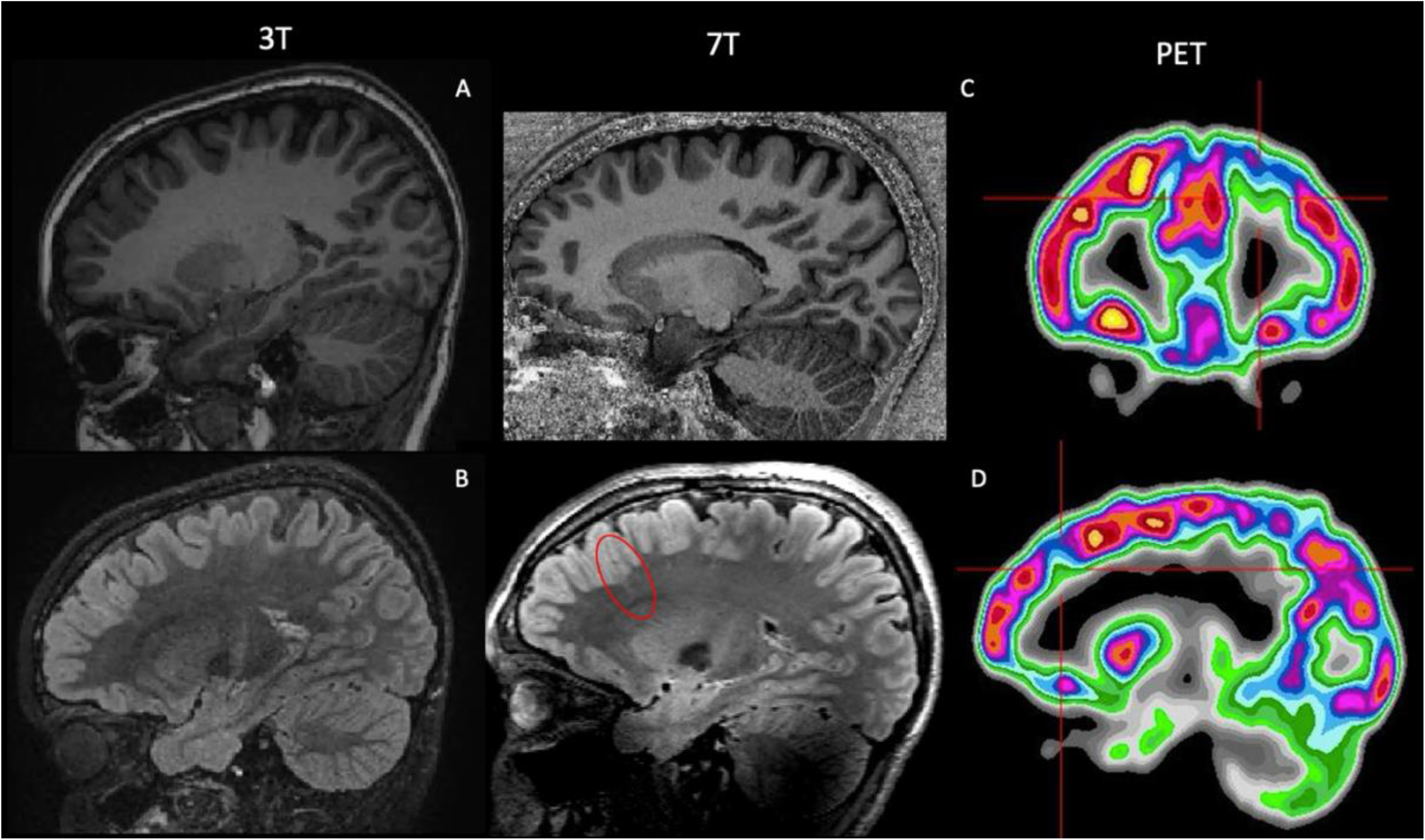
Case 1 (patient 1) Patient with severe treatment-resistant frontal lobe epilepsy, with nocturnal hypermotor seizures up to 3 times per week. The EEG suggested a left frontal lobe origin of seizures. The 3T MRI (A, B) highlighted the presence of a deeper left superior frontal sulcus but without an obvious cortical thickening or signal change in the grey-white matter interface. A cortical lesion was not seen. However, the PET scan (right) demonstrated heterogeneity of tracer uptake in the left frontal lobe compared to the right and a clear focal area of hypometabolism in the left superior frontal sulcus (clearly visible after co-registration). The 7T MP2RAGE (C) image did not show any abnormality, but the FLAIR (D) displayed disruption of the grey-white matter boundary and the presence of a subtle transmantle sign. The 7T FLAIR image matched the PET findings and correlated well with EEG and semiology. An FCD was suspected and because of the high conspicuity in an area of non-eloquent cortex, it was decided that there was no need for a stereo-EEG and the patient underwent neurosurgery. The histological finding was of an FCD type IIB. The patient was seizure free at 3 years follow-up.

**Figure 5:**
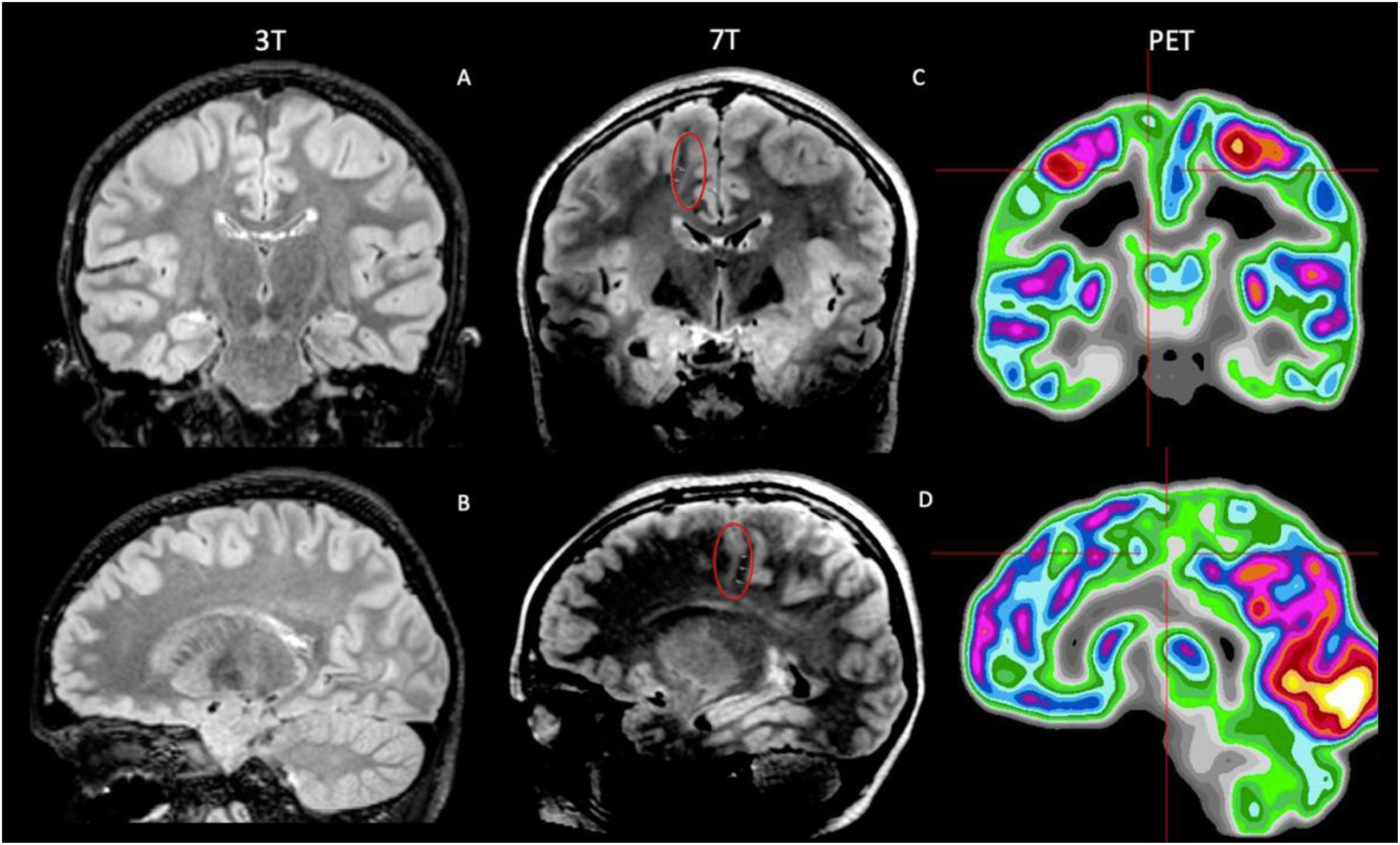
Case 2 (patient 7) Patient affected by severe frontal lobe epilepsy, with nocturnal seizures (up to 20 episodes per night). The EEG was consistent with structural epilepsy arising from the right frontal lobe, however, the 3T MRI (A, B) did not show a convincing lesion. The PET scan (right) detected a reduced tracer uptake in the right postcentral sulcus and right frontotemporal operculum. The 7T MRI (C, D) highlighted a structural abnormality originating in the right frontal lobe: a hyperintense signal extending from the medial frontal lobe cingulate gyrus area towards the lateral ventricle that was associated with ill-defined grey-white matter differentiation. This new information helped with restricting the area under investigation during stereo-EEG. The patient subsequently underwent surgery, and histological examination confirmed the presence of an FCD type IIB. The patient was seizure free at 3 years follow-up.

**Figure 6:**
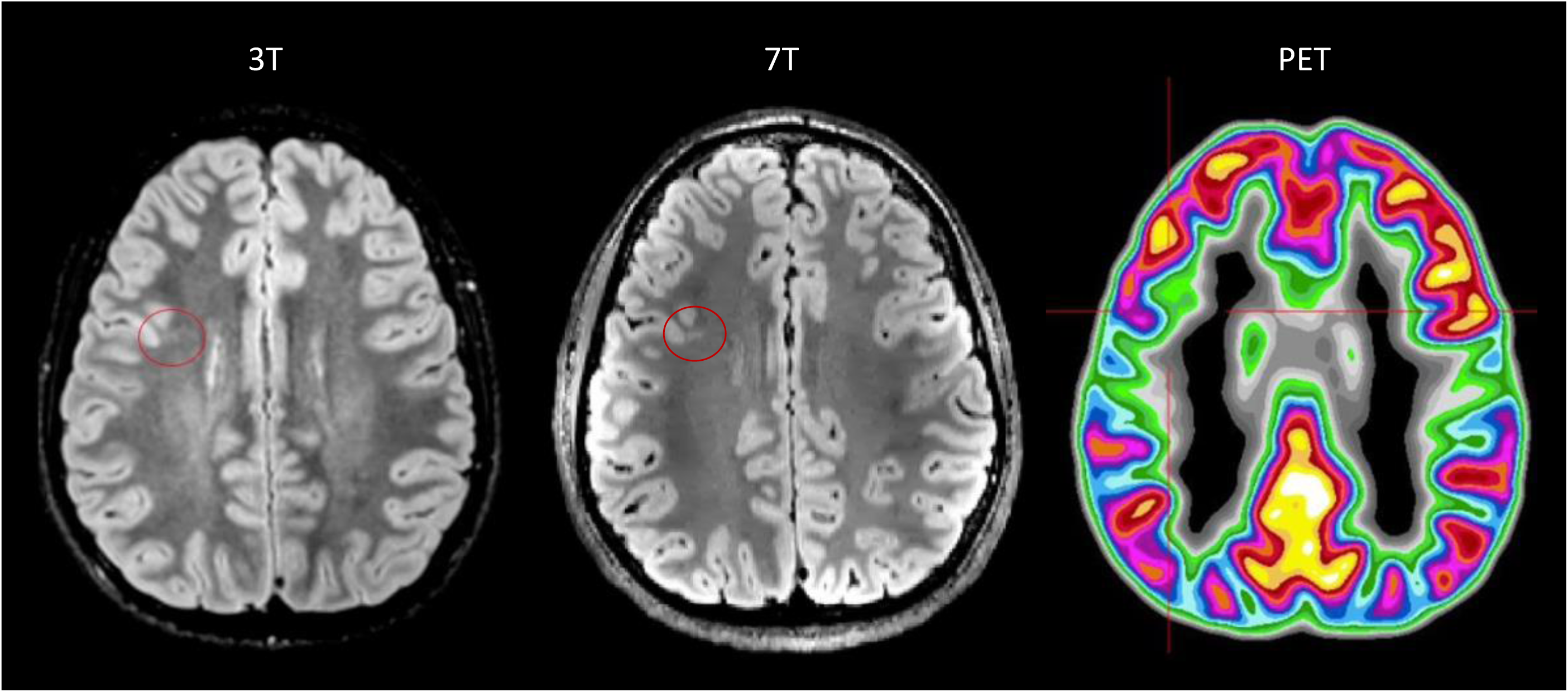
Case 3 (patient 18) Patient affected by predominantly nocturnal seizures (up to 4/5 times per week). The video-EEG captured seizures with electrical correlate from right frontal region, however, the 3T MRI (left) was reported as normal. The PET scan (right) detected a very focal reduced tracer uptake in the right inferior frontal sulcus. The 7T MRI (middle) highlighted a faint band of mildly increased signal in the FLAIR image extending into the white matter from the deep cortical margin in the depth of the right inferior frontal sulcus posteriorly (transmantle sign). This was detected precisely in the same sulcus as seen in the PET-MRI co registration. The stereo-EEG confirmed epileptic activity originating from that area. A thermocoagulation procedure was undertaken and the patient was seizure-free at 2 years follow up.

In patients that were lesion-positive at 3T (n=14), findings were replicated at 7T in all but one patient (patient 38) (93%). For this patient, polymicrogyria and an abnormal sulcus in the right middle frontal gyrus was suspected at 3T, which was concordant with the clinical EEG localisation. This finding was not confirmed at 7T.

There was one incidental finding at 7T in a 3T lesion-negative epileptic patient: a small cyst in the subcortical white matter of no clinical significance and unrelated to epilepsy. There were no new incidental findings at 7T MRI in the healthy control population.

## Discussion

We performed a systematic assessment of image quality, scan tolerability and clinical utility of 7T MRI scanning in childhood and in paediatric epilepsy. Overall, our work suggests that 7T MRI is both feasible and well-tolerated in children and has the potential to enhance lesion detection and aid pre-surgical planning in focal epilepsy cases with inconclusive conventional MRI results.

A key factor in understanding the practical utility of 7T MRI in children is to characterize the type and frequency of its associated side effects. Previous adult studies have shown that short-term effects such as dizziness, vertigo, nausea, headache and a metallic taste, may be experienced by subjects during ultra-high field scanning and that reported side-effects are larger than for lower field strengths (26,27,41–44). In line with this, our paediatric study demonstrated that increased dizziness was experienced during 7T scanning in comparison to 3T scanning. However, this settled after the subject had stopped moving through the field, none of the reported side effects persisted beyond the scanning procedure, and no adverse events were observed during the scanning session. Notably, side effects were more commonly reported amongst younger children, suggesting that the experience of higher field might differ based on age-related factors and developmental stages. This underscores the importance of tailoring procedures and providing adequate support and reassurance, especially for younger children.

Next, we investigated quantitative measures of image quality. Specifically, we compared cortical thickness measurements at 3T and 7T as a quantitative measure of a clinically relevant image characteristic. Our findings demonstrated thinner cortical boundaries at 7T, particularly in temporo-parietal/sensory regions. These results align with evidence from a previous study in adult participants (45), and suggest improved grey matter segmentation due to higher resolution an better contrast at 7T. Though an indirect measure (and not a ground truth of cortical thickness in the brain), the metric is a good proxy for grey / white matter boundary sharpness, the breakdown of which is a subtle marker of some cortical dysplasias.

Finally, clinical research in adults has demonstrated the sensitivity and clinical value of 7T MRI in patients with drug-resistant focal epilepsy, with a notable improvement in the detection rate of epileptogenic lesions ranging between 20 and 30% in those considered MRI negative at 3T (21–24). Here, 7T MRI revealed subtle lesions or suspicious areas in 31% of children with previously unrevealing 3T scans, mirroring the rates reported in adult populations. This underscores the potential of 7T as a valuable tool for improved diagnostic outcomes in paediatric focal epilepsy management.

These results should be considered a baseline. Wang et al. (40) highlighted the importance of post-processing approaches in increasing total lesion detection yield. Specifically, their study demonstrated that while unaided visual review alone generated 22% yield in 3T MRI negative patients, the addition of post-processing methods markedly increased yield to 43%. Additionally, the application of machine learning algorithms shows further promise in enhancing lesion detection in focal epilepsy. For example, the Multi-centre Epilepsy Lesion Detection (MELD) Project (46) demonstrated enhanced detection and classification of lesions in patients with focal epilepsy using an AI-driven approach, achieving a detection rate of 70% overall and 62% in MRI-negative cases.

In the present study, no new findings were reported in 3T MRI positive patients and typically developing controls. However, in one patient, polymicrogyria and an abnormal sulcus matching the interictal EEG abnormalities were suspected at 3T but were not confirmed at 7T. This discrepancy underscores the potential of 7T MRI’s higher resolution and sensitivity in potentially reducing false positives (40). Although it is essential to consider other factors such as imaging artifacts, which could have contributed to the contrasting results, no artefacts were reported in this patient’s scan.

Two of the eight patients with new findings at 7T have undergone surgical resection so far, and FCD type IIb were confirmed by histopathology for both. Because of the 7T findings confirming the PET-MR co-registration findings in these previously 3T-negative cases, stereo-EEG was re-planned with a reduced number of electrodes in one patient and omitted in the other. At three years after surgery, both patients were seizure-free. Another patient has undergone successful thermocoagulation after a subtle lesion was identified at 7T, confirming the PET-MR co-registration result, and was seizure-free at two years follow-up. In a fourth patient, the 7T scan helped with SEEG implantation and to better identify lesion margins. 7T may therefore be particularly useful when expert PET-MR co-registration is not available and has the potential of avoiding SEEG in selected cases.

This study has some limitations. First, patients had been extensively investigated in the Children’s Epilepsy Surgery Service (CESS) to precisely define the seizure-onset zone and decide patients’ management. This could have increased the probability of detecting structural abnormalities in our cohort. Second, 3T motion-corrected images were compared with 7T uncorrected images. In examinations impacted by motion, this might have prevented the visualisation of a structural lesion at 7T. Finally, not all patients had the 7T FLAIR acquired with parallel transmit as this had not yet been implemented when the study started. Therefore, artefacts due to B_1_ field inhomogeneities, particularly in the antero-basal right temporal lobe, could have impacted the detection of new findings at 7T in some cases. Nevertheless, 4/8 patients (50%) with new findings at 7T (patients 18, 24, 34, 41) were scanned with single transmit, suggesting that the improved detection at 7T is not solely dependent on the use of parallel transmit.

Despite these limitations, our findings suggest that 7T MRI in paediatric imaging is feasible, well-tolerated and associated with increased lesion-detection in children with focal epilepsy with unrevealing conventional MRI. 7T MRI may play an important role in facilitating pre-surgical planning and permitting a more focused and less invasive approach in these patients, leading to a better surgical outcome.

Importantly, future advancement in MR technology, such as the integration of motion correction and the utilization of advanced computational post-processing methods, have the potential to enhance the clinical value of 7T imaging in detecting structural brain lesions. Additionally, given the higher resolution and increased data volume produced by 7T MRI, any assistance in streamlining visual review processes is likely to yield substantial advantages. In particular, co-registration of PET and high-resolution MRI can provide an important prior to a focal hypothesis, thus facilitating the identification of the epileptogenic onset zone (18). Accordingly, in this study, 6/8 (75%) 3T MRI negative patients with new findings at 7T had prior PET findings.

Overall, this also highlights the importance of careful and appropriate selection of patients that are likely to benefit from a 7T investigation. Specifically, patients for whom a very specific pre-imaging hypothesis has been derived, from a combination of clinical and imaging data, may be the most likely to benefit from an ultra-high-resolution scan at 7T.

## Acknowledgements

We are grateful to the families who generously supported this study. We also thank the Paediatric Neurology team from Evelina London Children Hospital, including Dr. Ruth Williams, Dr. Elaine Hughes, Dr. Shan Tang, and Dr. Karine Lascelles.

## Supplementary Material

**Supplementary Table 1.** Clinical characteristics of the recruited patients. For completeness all patients scanned are described, however patient 16 was excluded from the final analysis due to extensive brain injury. In the last column, we report available clinical outcome updates for patients. Abbreviations: VNS: Vagus Nerve Stimulator; SEEG: Stereoelectroencephalography; WHO: World Health Organization.

| Patient | Sex | Disease duration (years) | EEG localization | PET localization | 3T MRI localisation | 7T localisation | Clinical-outcome updates |
| --- | --- | --- | --- | --- | --- | --- | --- |
| 1 | M | 7 | Left frontal | Left frontal | Negative | Left frontal | Left frontal craniotomy and resection of left superior frontal sulcus FCD with stealth and electrocorticography guidance. Seizure-free at 3 years follow-up. |
| 2 | M | 1 | Left occipital | Left temporal | Negative | Non-specific | Seizures became adequately controlled without surgery. |
| 3 | F | 8 | Left frontal | Left frontal | Negative | Negative | Seizures became adequately controlled without surgery. |
| 4 | M | 4 | Left frontal | Negative | Negative | Negative | VNS surgery. Good seizure control at three years follow-up. |
| 5 | F | 6 | Left temporal | Left temporal | Negative | Left temporal | VNS surgery. Not seizure-free. |
| 6 | F | 6 | Left parietal | N/A | Left parietal | Left parietal | SEEG and thermocoagulation. |
| 7 | M | 5 | Right frontal | Bilateral parietal | Negative | Right frontal | Right frontal convexity craniotomy for excision of a right supplementary motor area FCD. Seizure-free at 3 years follow-up. |
| 8 | F | 5 | Left occipital | Left parietal, left occipital | Negative | Negative | VNS surgery. VNS switched off due to poor tolerance. Not seizure-free. |
| 9 | M | 1 | Right parietal, right occipital | N/A | Right occipital | Right occipital | VNS surgery. Not seizure-free. |
| 10 | M | 2 | Right parietal | Right frontal | Right parietal | Right parietal | VNS surgery. |
| 11 | M | 3 | Left frontal, left temporal | N/A | Left frontal | Left frontal | Resection of left frontal tumour (Histology: Angiocentric glioma, WHO grade 1). Overall positive progress following surgery, but reports episodes of dizziness, breathlessness, tingling of the lips and blurred vision. |
| 12 | M | 5 | Right temporal | Right temporal | Negative | Negative | Seizures became adequately controlled without surgery. |
| 13 | M | 7 | Left temporal | Left temporal or left insula | Negative | Negative | Left temporal craniotomy. Awaiting pathology report. Seizure-free at 3 months follow-up. |
| 14 | F | 5 | Bilateral frontal and temporal | Left frontal, left temporal | Negative | Negative | N/A |
| 15 | F | 1 | Left temporal | Left temporal | Left temporal | Left temporal | Trying ketogenic diet. |
| 16 | M | 2 | Left frontal | N/A | Bilateral brain injury | Bilateral brain injury | Not a candidate for surgery. |
| 17 | F | 3 | Non-localising | N/A | Negative | Negative | N/A |
| 18 | M | 10 | Right frontal | Right frontal | Negative | Right frontal | SEEG and thermocoagulation. Seizure-free at two years follow-up. |
| 19 | F | 5 | Left temporal | Left temporal | Negative | Negative | N/A |
| 20 | F | 1 | Left frontal and temporal | N/A | Right parietal | Right parietal | Resection of right parietal tumour (Histology: Glioneuronal tumour, favouring ganglioglioma, WHO grade 1). Seizure-free at 2 years follow-up. |
| 21 | M | 8 | Left frontal and right frontal | Left superior temporal | Negative | Negative | SEEG revealing extensive bilateral involvement. Not suitable for surgery. |
| 22 | F | 14 | Non-localising | Left temporal and parietal | Left temporal | Left temporal | Left amygdalohippocampectomy showing mild reactive astrocytosis and possible neuronal loss in CA4 of the hippocampus. Seizure-free at 1 year follow-up. |
| 23 | F | 4 | Left hemisphere | Bilateral, multifocal | Negative | Negative | Pending further investigations. |
| 24 | F | 8 | Right frontal and temporal | Bilateral, multifocal | Negative | Not specific | Pending further MRI. |
| 25 | F | 2 | Left frontal and parietal | Left frontal & temporal | Negative | Negative | N/A |
| 26 | M | 10 | Left occipital and right occipital | Left frontal & temporal | Negative | Negative | Not a candidate for surgery |
| 27 | M | 0 | Left and right occipital | N/A | Negative | Negative | Not a candidate for surgery |
| 28 | M | 12 | Right temporal | Right temporal | Right temporal | Right temporal | N/A |
| 29 | F | 5 | Non-localising | Left parietal | Negative | Negative | Offered SEEG |
| 30 | F | 7 | Non-localising | N/A | Left hippocampal sclerosis | Left hippocampal sclerosis | Pending decision for surgery. |
| 31 | F | 2 | Left frontal | Left frontal | Negative | Negative | N/A |
| 32 | F | 9 | Non-localising | Left parietal | Negative | Negative | Not a candidate for surgery. |
| 33 | F | 2 | Posterior left hemisphere | N/A | Left temporal | Left temporal | Pending PET scan review. |
| 34 | F | 8 | Left temporal | Left temporal or left insula | Negative | Left temporal | N/A |
| 35 | M | 1 | Left parietal and temporal | N/A | Negative | Negative | N/A |
| 36 | M | 2 | Left frontal | N/A | Left frontal | Left frontal | Suspected FCD in the left superior frontal gyrus. 7T MRI helped to better identify lesion margins for SEEG implantation. On waiting list for surgery. |
| 37 | M | 2 | Right peri-rolandic region | N/A | Right perirolandic region | Right perirolandic region | Pending further investigations. |
| 38 | M | 3 | Right frontal | N/A | Right middle frontal gyrus | Negative | N/A |
| 39 | M | 6 | Right temporal and frontal | N/A | Negative | Negative | N/A |
| 40 | M | 6 | Left insula | N/A | Left insula | Left insula | N/A |
| 41 | M | 8 | Left frontal | N/A |  | Left frontal lobe | N/A |

**Supplementary Figure 1.**
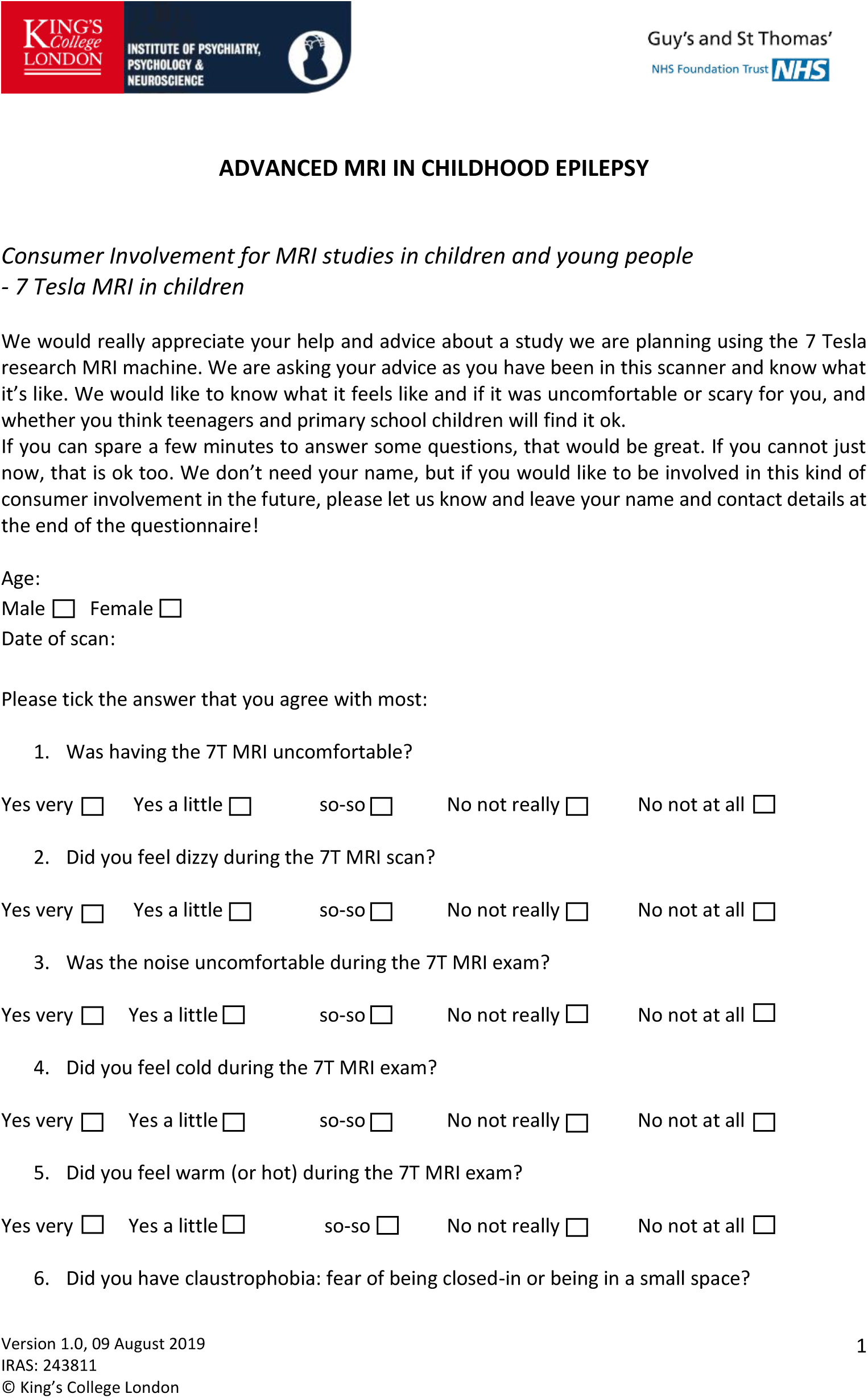
Safety and Tolerability Survey adapted from Chou et al. (34) employed to investigate general comfort and the presence of temporary collateral effects during 7T scanning.

